# Early health technology assessment of digital diabetes screening in Switzerland: cost-effectiveness and budget impact analyses

**DOI:** 10.64898/2026.02.10.26345992

**Authors:** Wasu Mekniran, Victoria Brügger, Magdalena Fuchs, Qiuhan Jin, Benjamin Wirth, Stefan Bilz, Michael Brändle, Elgar Fleisch, Tobias Kowatsch, Mia Jovanova

**Affiliations:** Department of Management, Technology and Economics, ETH Zurich, Zurich, Switzerland; Institute of Technology Management, University of St.Gallen, St.Gallen, Switzerland; School of Medicine, University of St.Gallen, St.Gallen, Switzerland; Institute for Implementation Science in Health Care, University of Zurich, Zurich, Switzerland; Department of Endocrinology, HOCH Health Ostschweiz, St.Gallen, Switzerland; Department of Internal Medicine, HOCH Health Ostschweiz, St.Gallen, Switzerland

**Keywords:** cost-effectiveness analysis, budget impact analysis, type 2 diabetes, digital biomarkers, Switzerland, payer incentives

## Abstract

Digital biomarkers offer scalable screening for type 2 diabetes, yet adoption is stalled by uncertainty regarding economic viability. This study evaluates the cost-effectiveness and budget impact of digital screening compared to opportunistic screening from a Swiss payer perspective.

**Methods:** A probabilistic Markov cohort model was developed to simulate at-risk Swiss adults (age ≥45, BMI ≥25 kg/m²) over a 40-year horizon. The model incorporates a digital attrition parameter, inputs derived from Swiss-specific sources (e.g., the CoLaus study and FSO life tables), and statutory tariffs. Costs and outcomes were discounted at 3.0%.

**Results:** In the deterministic base-case, digital screening yielded an incremental cost-effectiveness ratio of CHF 2,912 per quality-adjusted life-year gained. Probabilistic sensitivity analysis indicated a 93.2% probability of cost-effectiveness at the CHF 50,000 threshold. The budget impact analysis estimated a Year 1 gross investment budget of CHF 27 million to identify prevalent cases, followed by long-term savings from averted complications.

**Conclusions:** Digital screening can be highly cost-effective in Switzerland. While the required Year 1 gross investment poses a liquidity challenge, reimbursement via pathway-oriented models under the Swiss tariff could align incentives with long-term complication avoidance.

## 1. Introduction

Type 2 diabetes (T2D) affects 6% of the overall Swiss population and more than 12% in older age groups (≥45 years old), yet up to one-third of cases remain undiagnosed, shifting expenditure toward high-cost complications(1–3). The current standard of care, “opportunistic screening” for T2D, is structurally inefficient, constrained by high labor costs, limited access to general practitioners (GPs), and variable adherence to clinical practice guidelines(4–6). As a result, screening coverage is limited, and case-finding is inconsistent(7).

Digital risk biomarkers in this context refer to objective, consumer-generated physiological signals, such as photoplethysmography (PPG) via a smartphone camera (8,9) or voice acoustics patterns via a smartphone microphone, that serve as scalable proxies for glycemic risk(10). By decoupling triage from clinics, these tools theoretically lower the marginal cost of screening, even compared with traditional community-based programs (6,11,12). However, technological scalability does not guarantee economic viability. For statutory health insurers, adopting these digital screening tools requires crossing an intertemporal trade-off: accepting an immediate surge in diagnostic expenditures to secure long-term savings from averted complications(1). Insufficient specificity (8) may increase unnecessary downstream confirmatory testing and consultation costs(12). This effectiveness estimate is further complicated by the transition of the outpatient tariff from TARMED to bundled TARDOC, which recalibrates Swiss outpatient reimbursement and prioritizes intellectual medical performance over technical interventions. At the same time, this shift increases the unit cost of physician-led screening, necessitating periodic regulatory revision cycles to keep cost in balance. Crucially, the introduction of dedicated tariff positions for digital services formalizes the revenue for remote prevention, addressing previously unbilled coordination into a measurable fiscal constraint within the healthcare system’s broader incentive structure(13).

To align healthcare digitalization with payer reimbursement standards, this study applies a development-focused health technology assessment (HTA) framework(14–17). We constructed a six-state probabilistic Markov model (healthy, prediabetes, undiagnosed T2D, diagnosed T2D, complications, death) to simulate the lifetime clinical and economic trajectory of the Swiss at-risk adult cohort. Rather than assuming ideal implementation, we modeled the viability frontier of these tools, explicitly accounting for real-world friction such as imperfect algorithmic accuracy(8) and patient non-compliance with confirmatory tests following screening. Accordingly, this analysis addresses two research questions (RQs): (RQ1) Is digital screening cost-effective relative to opportunistic screening for T2D? (RQ2) How does national implementation of digital screening affect short-term payer budgets and long-term complication-related expenditures?

## 2. Methods

### 2.1 Study design and model framework

To answer both RQ1 and RQ2, we performed a cost-effectiveness analysis (CEA) (18) and budget impact analysis (BIA) (19), assessing the economic feasibility of digital screening for T2D(20–22). The study adhered to the Consolidated Health Economic Evaluation Reporting Standards (CHEERS 2022) (23). We developed a six-state probabilistic Markov model to simulate a closed synthetic cohort of 1.54 million Swiss adults, defined as individuals aged ≥45 with a BMI ≥25 kg/m²; this cohort size was derived by applying age- and BMI-specific prevalence rates from the Federal Statistical Office(24). This model excludes new entrants over the time horizon, tracking only the initial population until the study conclusion, isolating longitudinal progression. The fixed denominator model uses a 1-year cycle length, applying half-cycle corrections to both costs and outcomes to account for transitions occurring mid-cycle(4,5,25). The simulation was constructed using Python 3.13 (NumPy v1.20 for matrix operations, Pandas v1.3 for data management, matplotlib v3.4, and seaborn v0.11 for visualization). To ensure reproducibility, a random seed was set for all 1,000 Monte Carlo iterations.

The analysis employs a 40-year lifetime horizon to fully capture the longitudinal benefits of early dysglycemia detection, and the averted costs of late-stage complications, as age is one of the strongest risk factors for T2D, based on data published by the DECODE study group(26). A secondary 5-year horizon was used for the BIA (19) to align with payer budget cycles and the TARDOC tariff renegotiation windows. Future costs and health outcomes, measured in quality-adjusted life years (QALYs), were discounted at an annual rate of 3.0%, consistent with Swiss health economic guidelines(18,23). Since this early-stage, development-focused HTA aimed to inform pre-market parameterization rather than evaluate a historical intervention, a formal health economic analysis plan was not pre-registered. Instead, the analysis was conducted using an iterative simulation approach.

All variable assumptions are reported in Table 1. These values represent modelled assumptions based on publications and public sources; no proprietary company or personal health data were used. The full uncertainty program, including Monte Carlo simulation, one-way sensitivity analysis, and two-way elasticity analysis, along with all underlying code and reproducible scripts, is available in the public repository (DOI: <u>10.17605/OSF.IO/5G2YR</u>)

**Table 1.** Detailed model parameters and distributional hyperparameters.

| Parameter category | Parameter name | Base case (mean) | Std. Error (SE)* | Distribution | Hyperparameters (calculated)** | Source |
| --- | --- | --- | --- | --- | --- | --- |
| Cohort & epidemiology | Diabetes prevalence (Age $\geq 45$) | 0.12 | 0.02 | Beta | $\alpha=31.5, \beta=231.5$ | (51) |
| | Prediabetes prevalence | 0.309 | 0.038 | Beta | $\alpha=42.6, \beta=95.4$ | (3) |
| | Undiagnosed diabetes fraction | 0.3 | 0.076 | Beta | $\alpha=10.2, \beta=23.8$ | (51) |
| | Overweight share | 0.45 | 0.025 | Beta | $\alpha=197.5, \beta=241.4$ | (24) |
| Transition probabilities | Prediabetes to T2D (SoC) | 0.06 | 0.008 | Beta | $\alpha=52.8, \beta=827.2$ | (50) |
| | Prediabetes to T2D (int) | 0.03 | 0.005 | Beta | $\alpha=34.9, \beta=1128.1$ | Modeled (-50% RR) |
| | Regression benefit (int add-on)*** | 0.015 | 0.005 | Beta | $\alpha=8.8, \beta=581.2$ | (50) |
| | T2D to Prediabetes | 0.005 | 0.002 | Beta | $\alpha=6.2, \beta=1238.8$ | Calibration |
| Diagnostic performance | Sensitivity (int) | 0.7 | 0.075 | Beta | $\alpha=19.6, \beta=8.4$ | (8) |
| | Specificity (int) | 0.7 | 0.075 | Beta | $\alpha=19.6, \beta=8.4$ | (8) |
| | Digital attrition (uptake rate) | 0.3 | 0.05 | Beta | $\alpha=24.9, \beta=58.1$ | (45) |
| | Smartphone access share | 0.85 | 0.025 | Beta | $\alpha=39.5, \beta=7.0$ | (24) |
| Costs (2025 CHF) | Screening unit cost (int) | 40 | 4 | Gamma | $k=100.0, \theta=0.40$ | (11) |
| | Blood glucose confirmatory test | 75 | 7.5 | Gamma | $k=100.0, \theta=0.75$ | (39) |
| | Initial treatment cost (GP workup) | 240 | 24 | Gamma | $k=100.0, \theta=2.40$ | (13) |
| | Prediabetes mgmt (SoC) | 150 | 15 | Gamma | $k=100.0, \theta=1.50$ | Assumption (GP) |
| | Prediabetes mgmt (intervention) | 480 | 48 | Gamma | $k=100.0, \theta=4.80$ | (50) |
| | Annual cost: diagnosed T2D | 3,324 | 332 | Gamma | $k=100.0, \theta=33.24$ | (52) |
| | Annual cost: complications | 5,500 | 550 | Gamma | $k=100.0, \theta=55.00$ | (1) |
| Utilities<br>(QALYs) | Utility:<br>prediabetes | 0.89 | 0.051 | Beta | $\alpha=16.8, \beta=2.1$ | (47) |
| | Utility:<br>diagnosed T2D | 0.7 | 0.051 | Beta | $\alpha=36.7, \beta=15.7$ | (48) |
| | Utility:<br>complications | 0.6 | 0.051 | Beta | $\alpha=47.4, \beta=31.6$ | (49) |

\*Note: SE estimated assuming 10%–20% coefficient of variation where explicit CIs
were unavailable. \*\*Note: Hyperparameters derived via Method of Moments. \*\*\*Note:
The model applies this 0.015 as an additive benefit to the natural regression rate
(0.0162), yielding a total regression probability of 0.0312 in the intervention arm.

### 2.2 Model structure and disease progression

The model comprises six health states: healthy (normoglycemia), prediabetes, undiagnosed T2D, diagnosed T2D, diabetes-related complications, and death, see Figure 1. Transitions between states are governed by time-dependent probabilities referred to the longitudinal epidemiological data from CoLaus study, ADDITION-Europe Study and the US Diabetes Prevention Program (27–30). The model distinguishes explicitly between undiagnosed and diagnosed T2D; while both states incur disease progression risks, only the diagnosed state incurs management costs and permits modeled treatment effects(31). All-cause mortality rates were modelled based on the Swiss Federal Statistical Office(32) life tables, with excess mortality hazards associated with T2D and its complications applied as multipliers, consistent with the significant years of potential life lost observed in long-term metabolic cohorts in previous studies(33–35).

**Figure 1.**
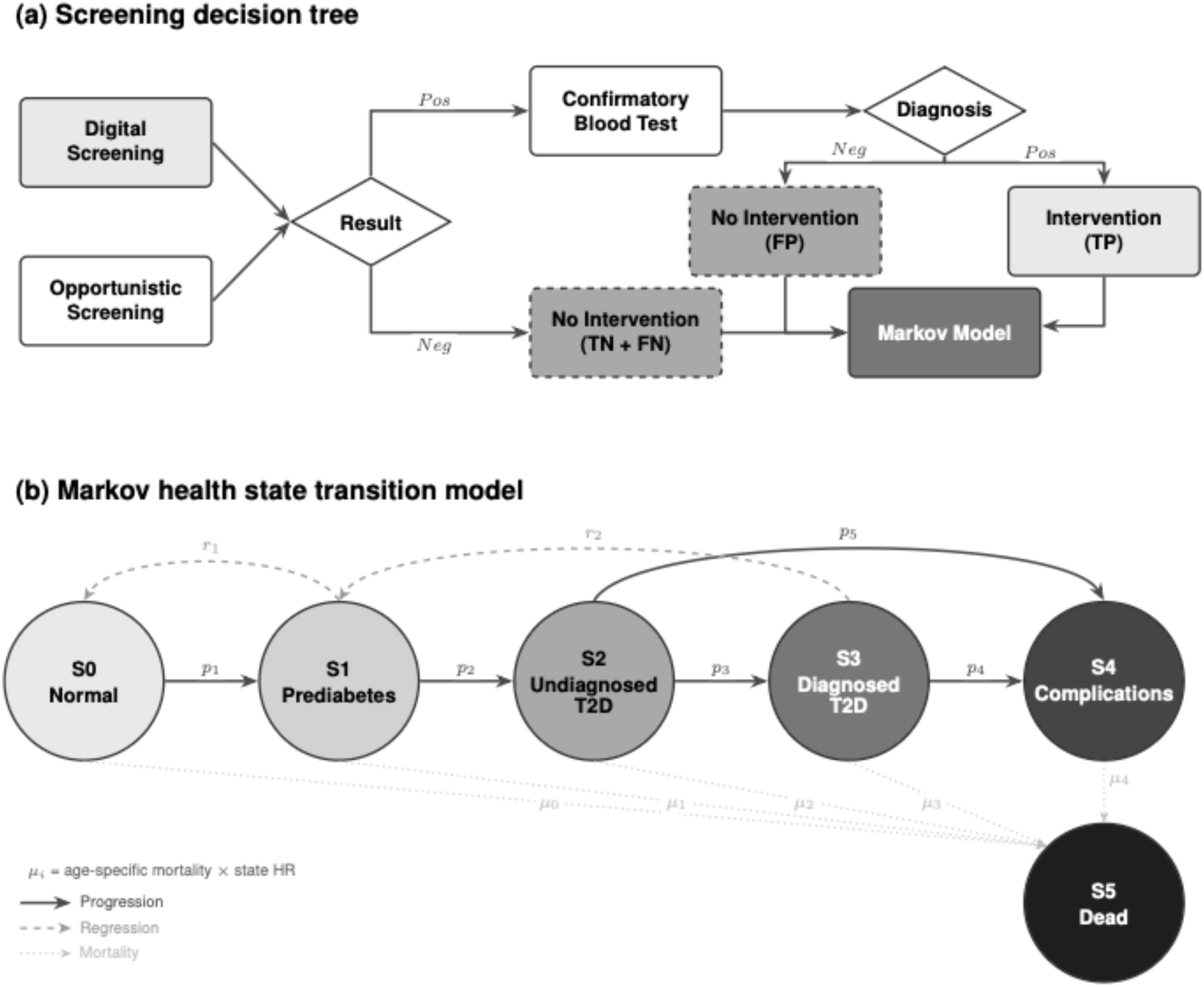
Model structure includes (a) screening decision tree and (b) Markov health state transition.

The analysis compares two primary screening pathways. The comparator arm reflects the current standard of care (SoC): opportunistic screening occurring stochastically during unrelated GP visits(12,31,36,37). We modelled an annual background diagnosis rate of approximately 10%(31) for the undiagnosed pool, calibrated to reflect periodic opportunistic screening in recommended routine checkups(4–6).

### 2.3 Data inputs and parameterization

The intervention arm simulates a two-step digital triage pathway, including a screening step and a confirmatory test step, see Figure 1(a). In the first step, individuals engage with a smartphone application for data collection and risk detection(8,36–38). Base-case parameters of the digital screening were modelled with a sensitivity and specificity of 70% each, with a unit cost of CHF 40 representing the full economic cost of software provision and data infrastructure(11,39). Individuals flagged as at-risk in the first screening step were referred for confirmatory blood glucose testing in the second step. A conservative cost of CHF 75 is set based on commercial home-test pricing, substantially higher than the statutory laboratory tariff (∼CHF 20) (13), ensuring our model overestimates rather than underestimates downstream diagnostic costs.

In the intervention arm, the annual progression rate was conservative at 3% annual progression rate (approx. 50% relative risk reduction vs. 58% in DPP study) to reflect real-world effectiveness(40,41). Regression from prediabetes to normoglycemia was parameterized using longitudinal data from ADDITION-Europe(29). In the standard-of-care arm, the annual reversion probability was set at 1.62%, derived from observed three-year follow-up data(42). In the intervention arm, this probability was increased to 3.12% annually, reflecting modest behavioral activation(27,43). It enhanced clinical engagement following risk identification, consistent with previous long-term findings.

To ensure reproducibility of a probabilistic sensitivity analysis (PSA), this table details the distributional assumptions, standard errors (SE), and calculated hyperparameters (α, β for Beta distributions; k, θ for Gamma distributions) used in the 1,000 Monte Carlo simulations. Standard errors were derived from reported 95% confidence intervals (CIs) or assumed to be 10%–20% of the mean when variance data were unavailable, consistent with ISPOR modelling good practices(19).

Real-world adoption friction was explicitly modelled through a digital attrition, the drop-off between risk signal and clinical confirmation, where only 30% of individuals flagged as at-risk complete confirmatory blood glucose testing(44). This assumption reflects the observed drop-off in digital risk notification and clinical follow-up in digital health interventions and prevents overestimating effective case detection(45).

Health outcomes were expressed in QALYs. Health state utility weights were drawn from established diabetes-specific valuation studies and assigned as follows: healthy (0.90) (46), prediabetes (0.89) (47), undiagnosed diabetes (0.70) (48), diagnosed diabetes without complications (0.70) (48), and diabetes with complications (0.60) (49). The model distinguishes between undiagnosed and diagnosed T2D states through a 50% higher hazard ratio for progression to complications in the undiagnosed state, reflecting the adverse metabolic consequences of delayed treatment initiation while avoiding assumptions about patient-experienced symptom burden prior to diagnosis. Although Swiss-specific utility estimates were unavailable, the selected values are consistent with prior cost-effectiveness analyses in diabetes(11,21,50) and were extensively tested in sensitivity analyses, see a Markov chain cohort trace in supplementary material Table S1.

### 2.4 Uncertainty and decision rules

To accurately reflect the stochastic properties of the model inputs, beta distributions were fitted to bounded probability parameters (transition rates, diagnostic accuracy, utility weights). In contrast, gamma distributions were applied to cost parameters to capture the right-skewed nature of healthcare expenditure data. The primary economic outcome was the incremental cost-effectiveness ratio (ICER). The digital screening tool was deemed cost-effective if the ICER fell below the willingness-to-pay (WTP) threshold of CHF 50,000 per QALY, consistent with stricter Swiss empirical benchmarks (53).

Similarly, the results were displayed using a cost-effectiveness acceptability curve to illustrate the likelihood that digital screening is cost-effective at various WTP thresholds over time. To quantify the value of resolving parameter uncertainty, we calculated the expected value of perfect information (EVPI) using standard methods(54). EVPI represents the maximum amount a decision-maker should be willing to pay to eliminate all parameter uncertainty before making the adoption decision.

For each willingness-to-pay threshold (λ = CHF 0 to 100,000 per QALY, in CHF 2,000 increments), EVPI was computed as:

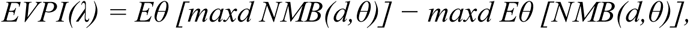

where NMB denotes net monetary benefit (λ × ΔQALYs − ΔCosts), d represents the decision (digital vs. opportunistic screening), and θ represents uncertain parameters sampled in the PSA. The first term reflects the expected benefit of always choosing the optimal strategy for each simulation (perfect information). In contrast, the second term reflects the expected benefit of committing to a single strategy based on average outcomes (current information). Per-person EVPI was calculated by dividing total NMB by the target population size. Population-level EVPI was subsequently estimated by multiplying per-person EVPI by the affected population (n = 4,042,032). These estimates represent an upper bound on the economic value of additional research, rather than prescriptive guidance for specific study designs(53–55). The findings of the cost-effectiveness and budget impact analyses are presented below, beginning with base-case results followed by sensitivity and scenario analyses.

## 3. Results

### 3.1 Base case cost-effectiveness and allocative efficiency (RQ1)

As detailed in Table 2, the deterministic base-case analysis yielded an ICER of CHF 2,912 per QALY gained. This efficiency compares favorably with existing economic evaluations of both traditional and digital prevention strategies(50,56).

**Table 2.** Base-case cost-effectiveness results (per capita and per person screened).

| Metric | Opportunistic screening | Digital screening | Incremental |
| --- | --- | --- | --- |
| Population analysis (per capita) |  |  |  |
| Screening test costs | 0.98 | 3.33 | 2.35 |
| Confirmatory test costs | 1.89 | 2.28 | 0.39 |
| Initial treatment costs | 1.62 | 2.27 | 0.65 |
| Long-term treatment costs | 13,069.93 | 13,102.42 | 32.49 |
| Complication costs | 6,012.97 | 5,999.82 | −13.15 |
| Total costs | 19,085.77 | 19,107.85 | 22.08 |
| Life years (undiscounted) | 31.006 | 31.022 | 0.016 |
| QALYs (discounted) | 16.42 | 16.427 | 0.008 |
| ICER (CHF per QALY) | - | 2,912 |  |
| Screened cohort analysis (per person) |  |  |  |
| Screening test costs | 20 | 40 | 20 |
| Confirmatory test costs | 38.61 | 27.36 | −11.25 |
| Initial treatment costs | 33.05 | 27.22 | −5.83 |
| Long-term treatment costs | 266,832.41 | 157,350.41 | −109,482.00 |
| Complication costs | 122,759.35 | 72,053.49 | −50,705.86 |
| **Total costs** | **389,650.37** | **229,471.26** | **−160,179.11** |
| QALYs gained | 53.138 | 53.163 | 0.025 |

### 3.2 Probabilistic sensitivity analysis (RQ1)

The robustness of CEA findings was confirmed through probabilistic sensitivity analysis (PSA), see Figure 2. Across 1,000 stochastic simulations over the 40-year lifetime horizon, the digital screening pathway yielded a median incremental cost of CHF 98 million (95% Credible Interval (CrI): –19.9 million to 285.2 million) and median incremental health gains of 26,119 QALYs (95% CrI: –6,548 to 79,819). These outcomes correspond to a base case ICER of CHF 3,958 per QALY gained. Consequently, the probability of cost-effectiveness at the CHF 50,000 threshold was 93%, indicating a high degree of decision certainty.

**Figure 2.**
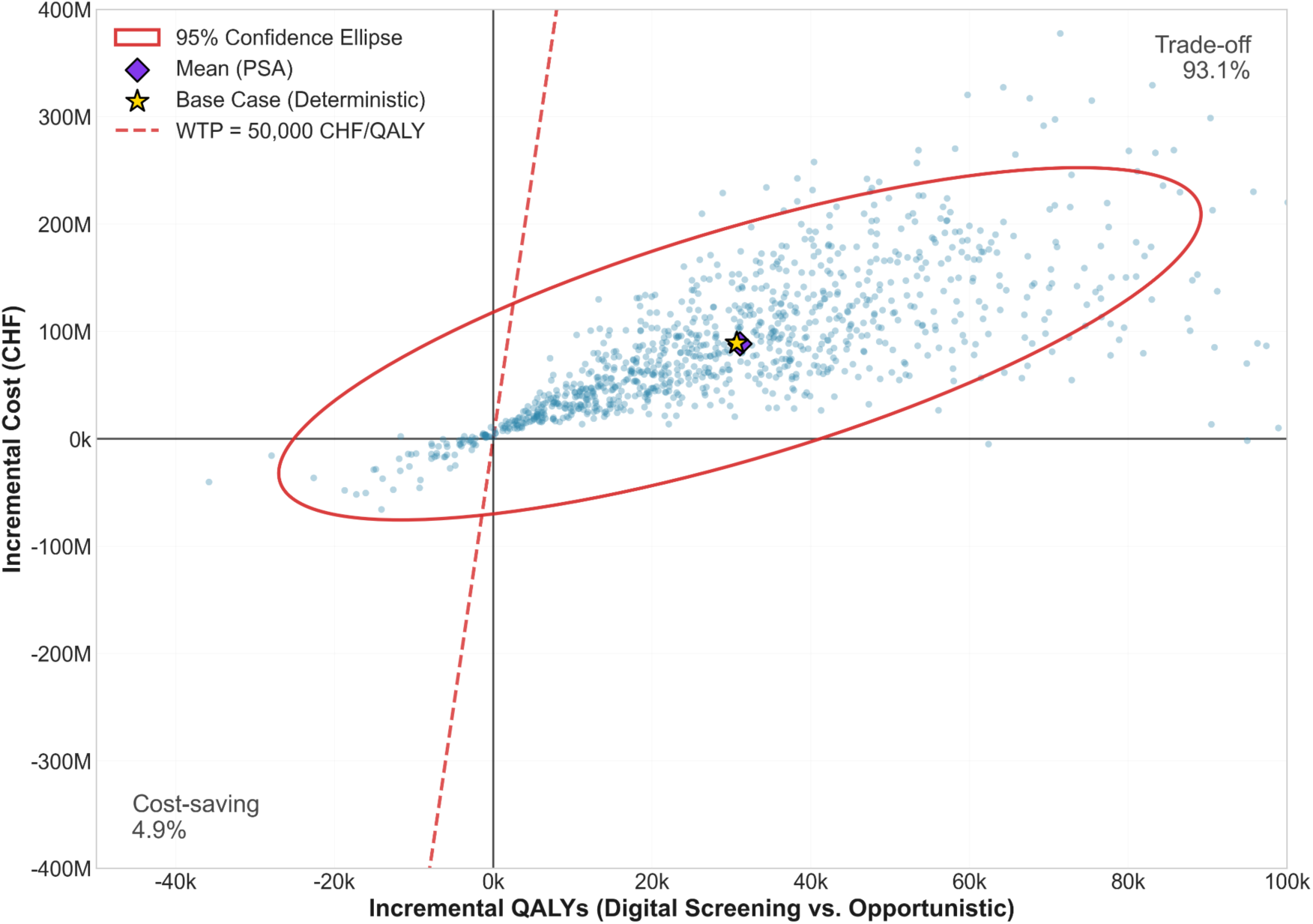
Cost-effectiveness plane (40-year horizon) shows that 93% of iterations fall within the cost-effective quadrant (north-east), with the upper bound remaining below the WTP threshold of CHF 50,000.

Decision uncertainty is further quantified in the cost-effectiveness acceptability curve (CEAC). The curve shows a rapid convergence to probability 1.0 at the standard threshold of CHF 50,000 per QALY, in supplementary material Figure S5. The probability of the digital screening being cost-effective at 93.2%, compared with 6.8% for opportunistic screening. Crucially, this probability remains robust even under stricter fiscal constraints. At a conservative threshold of CHF 15,000, the digital screening retains a high probability of cost-effectiveness, suggesting that the economic value is resilient to substantial parameter volatility and varying policy benchmarks.

### 3.3 Deterministic sensitivity and value of information analysis (RQ1)

#### One-way sensitivity analysis

Univariate deterministic sensitivity analysis identified the primary drivers of model uncertainty. As illustrated in the tornado diagram (Figure 3), the ICER was most sensitive to variations in the baseline prevalence of undiagnosed prediabetes and the unit cost of the screening. Secondary drivers included the costs of lifestyle intervention management and the underlying prevalence of prediabetes in the target cohort. In contrast, the inelasticity of the cost of confirmatory blood glucose testing is expected, as the same confirmatory diagnostic standard is used in both the digital screening and opportunistic pathways.

**Figure 3.**
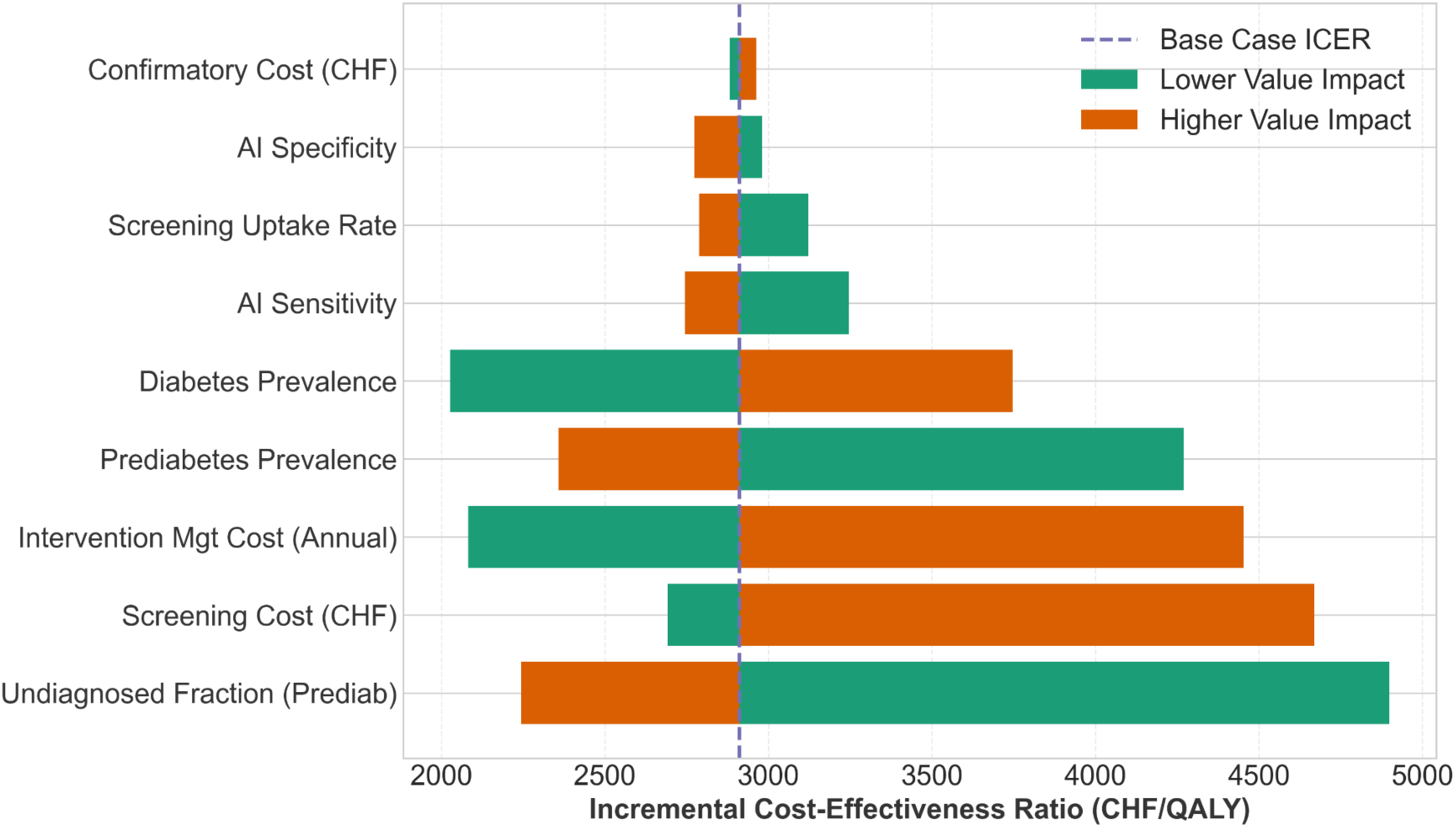
Tornado diagram of one-way sensitivity analysis. The ICER is most sensitive to the prevalence of undiagnosed prediabetes and screening cost. No single parameter variation causes the ICER to exceed the CHF 50,000 threshold, indicating deterministic robustness.

#### Threshold analysis with two-way sensitivity analysis

We distinguished between the actual modeled cost and the maximum allowable cost to maintain cost-effectiveness. In the base case, the digital screening unit cost was modeled at CHF 40. Threshold analysis demonstrates that the digital screening preserves substantial fiscal headroom: even at the upper bound of our analysis, a screening unit cost of CHF 200 per person, the ICER remains significantly below the CHF 50,000 WTP threshold, indicating a robust allowable price, shown in Figure 4.

**Figure 4.**
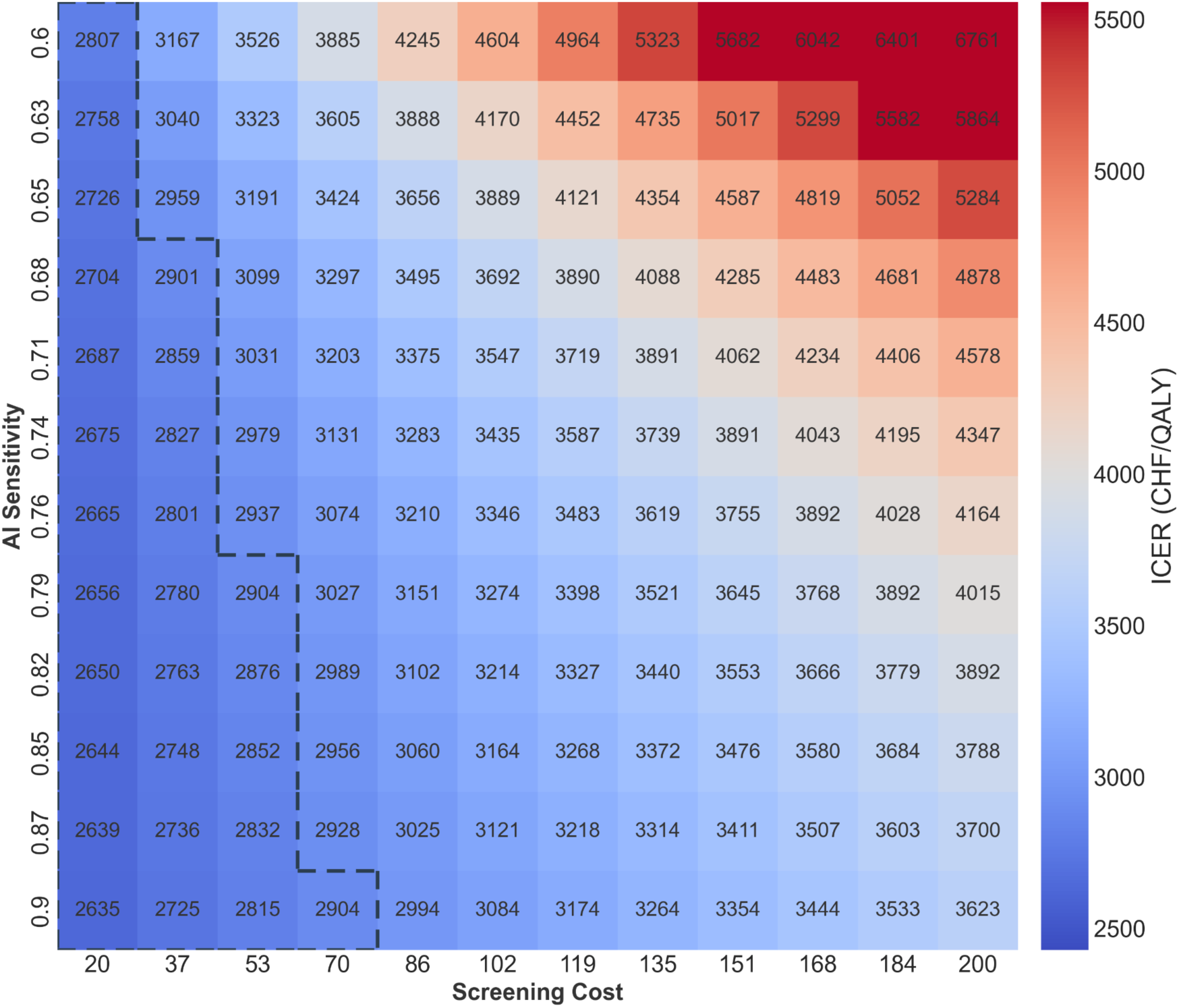
Threshold heatmap of ICER by sensitivity and cost.

Additionally, the dashed line marks the viability frontier based on the base-case ICER of CHF 2,912 per QALY. It shows that even if diagnostic sensitivity drops to 0.60 (leading to fewer detected cases), digital screening remains comparable to the base-case scenario, as long as the screening cost stays below CHF 37. Moreover, it can maintain the same level of cost-effectiveness up to a screening cost of CHF 70, if the sensitivity of 0.9 is achieved. Either way, the simulated range stays entirely below the Swiss WTP threshold.

### 3.4 Budget impact analysis (RQ2)

#### Short-term liquidity (t=1 to t=5)

The BIA reveals an intertemporal asymmetry between immediate liabilities and long-term savings. In Year 1, the nationwide rollout requires a gross upfront diagnostic investment of CHF 27 million to clear the historical backlog of undiagnosed disease, characterized as a prevalence tax encompassing the screening of a projected 336,649 individuals (30% uptake). Following this initial phase, costs stabilize as the program targets further incident cases. By Year 5, the cumulative net budget impact is CHF 59 million, as immediate screening costs are partially offset by CHF 8.8 million in averted complication costs.

The prevalence tax dynamic is visible. High upfront screening and treatment costs for prevalent cases (red bars) are partially offset within the first 6 years, resulting in a net budget impact of CHF 63 million, as a digital screening matched the cost-effectiveness of standard of care opportunistic screening, see the CEAC in Figure S4.

#### Long-Term liability reduction (t=40)

Over the 40-year simulation horizon starting at age 45 years, the cumulative net budget impact is projected at CHF 122 million. Crucially, the net budget impact is significantly mitigated by a delayed disease progression, accounting for a savings of CHF 48 million and a complication cost offset of CHF 53 million, see the waterfall cost decomposition in Figure S9. Age-stratified analysis reveals that the timing and magnitude of these returns are highly cohort-dependent: while absolute QALY gains are most significant for individuals aged 50–55 years (after Year 10 in simulation), the fiscal efficiency of the digital screening is most favorable in older groups, with the ICER reaching its lowest point of CHF 2,188 at age 70 years, see this effect in Figure 6. As the cohort ages, returns on early detection materialize through the suppression of capital-intensive morbidities.

**Figure 5.**
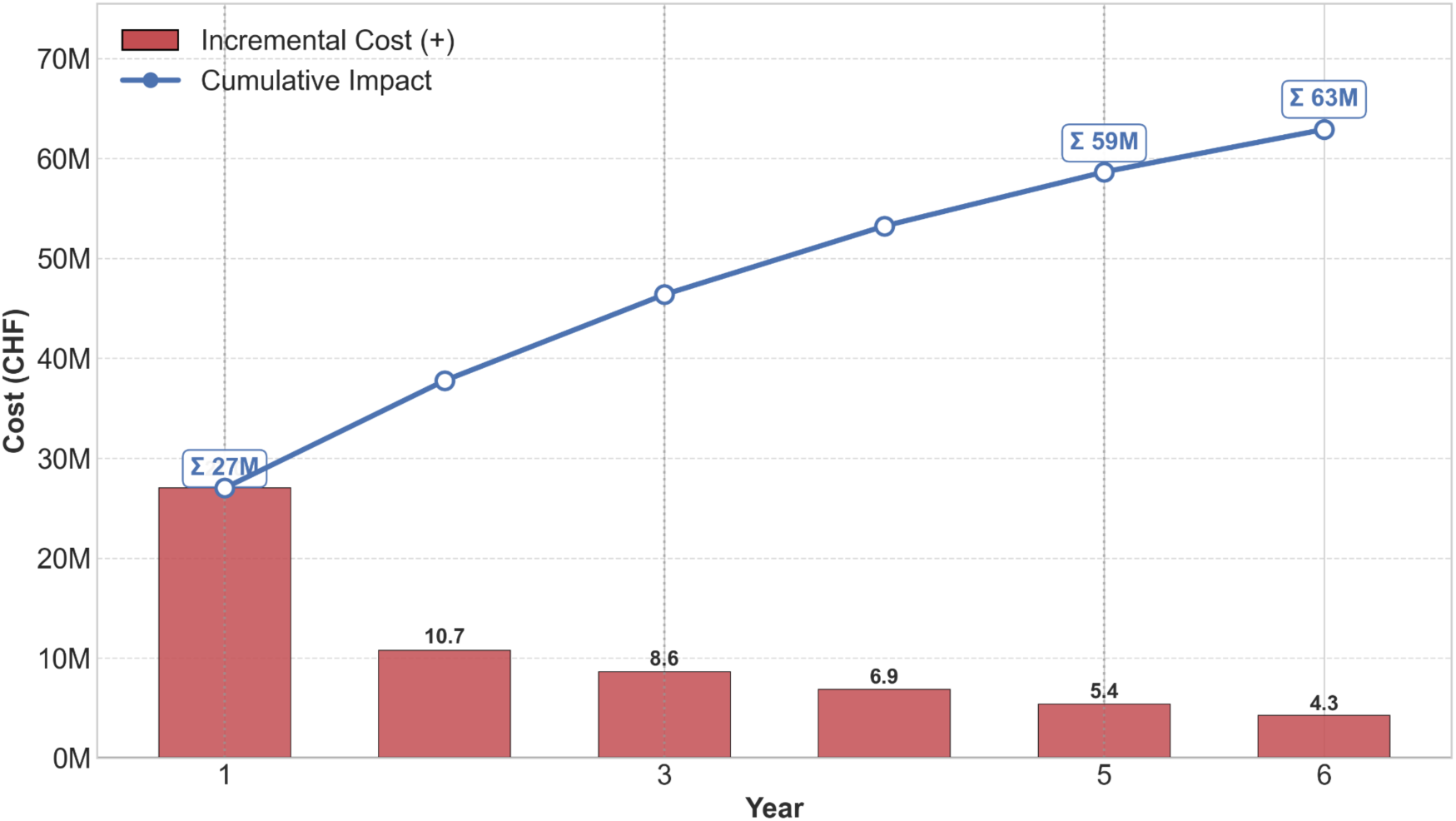
Budget impact decomposition in short-term horizon.

**Figure 6.**
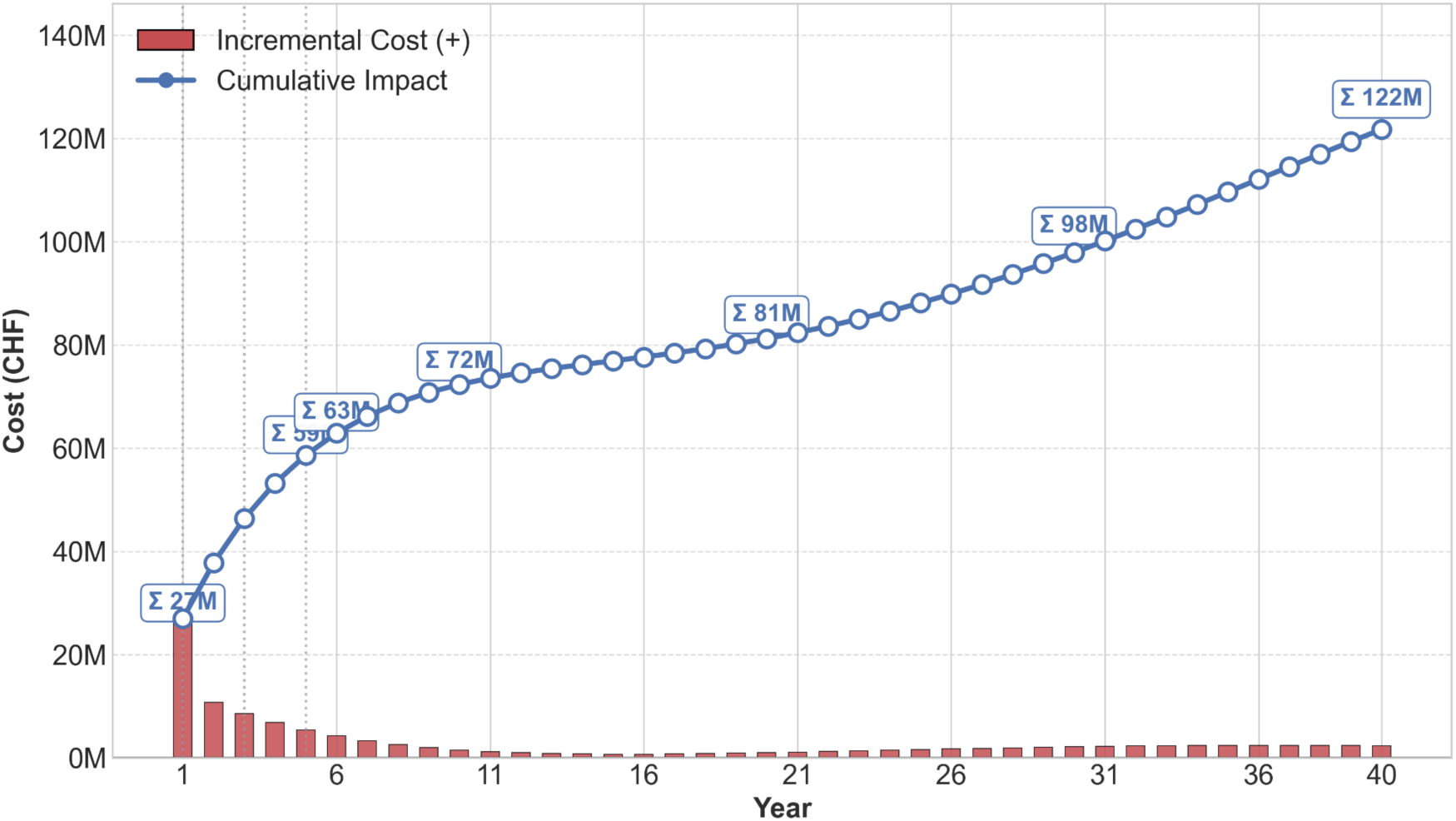
Annual budget impact of digital vs. opportunistic screening (40-year horizon). The cumulative budget impact stabilizes after the initial prevalence tax, with long-term cost curves flattening as complication savings offset maintenance screening costs.

#### Value of information

The EVPI analysis indicated that residual decision uncertainty has minimal impact on adoption decisions. At the WTP threshold of CHF 50,000 per QALY, per-person EVPI was CHF 5.19, reflecting the low probability (1.6%) that the digital screening strategy would be suboptimal. Population-level EVPI was approximately CHF 21.0 million over a 40-year lifetime horizon (equivalent to CHF 0.52 million per year). This value represents the maximum amount that could be economically justified for additional research aimed at eliminating remaining decision uncertainty.

## 4. Discussion

### 4.1 Principal findings

#### The economics of triage

Under the deterministic base-case assumptions, the ICER of CHF 2,912 per QALY demonstrates substantial economic value. This conclusion is supported by the PSA, which indicates a 93% likelihood of cost-effectiveness at the common Swiss standards(53). This conclusion is further supported by a narrow 95% credible interval (CHF 1,453–8,841), clearly indicating that economic inefficiency is unlikely.

While traditional screening in dental or community settings has yielded ICERs ranging from cost-saving to $9,511 per QALY(11,20), digital interventions often exhibit higher variability, with recent evaluations of prescription digital therapeutics and digital diabetes prevention programs (dDPP) reporting ICERs between $6,212 and $17,443 per QALY(57,58), with some studies indicating that digital formats can be cost-saving from a societal perspective(59,60).

Our result falls toward the more efficient end of this spectrum, a distinction likely driven by the structural differences between screening and treatment(56). Unlike dDPP models that often retain variable human-coaching costs, the modeled digital screening step relies on automated, low-marginal-cost triage to identify risk. This positions digital screening as a highly scalable ‘top-of-funnel’ mechanism distinct from downstream therapeutic costs. Consequently, the resulting ICER is substantially below the implicit Swiss WTP threshold of CHF 50,000 per QALY, indicating robust cost-effectiveness.

#### Decoupling accuracy from value

Our findings indicate that achieving cost-effectiveness in a triage context does not require near-perfect diagnostic specificity(8). Our threshold analysis shows that a screening tool with moderate specificity of 70% can remain financially viable because its low marginal cost (CHF 40.00) allows sufficient budget to cover subsequent confirmatory tests (CHF 75.00) (39) for false positive cases. As demonstrated by our sensitivity analyses, the economic viability of digital screening relies on balancing the screening unit cost with the downstream costs of missed cases, rather than requiring nearly perfect accuracy. Low-cost screening broadens access to at-risk populations that are less likely to be reached through labor-intensive opportunistic screening (6), thus enhancing case detection without sacrificing cost-effectiveness (61).

To contextualize these estimates, a prospective validation study of digital diabetes screening algorithms in Swiss primary care settings would typically cost CHF 2–5 million, depending on sample size and follow-up duration(62). A pragmatic cluster-randomized trial comparing screening strategies would require CHF 8–15 million(41). Given that the population-level EVPI of CHF 21 million represents the maximum value of eliminating all parameter uncertainty, the economic case for delaying implementation pending additional evidence is considered weak.

#### Robustness and equity implications

The economic case for digital screening remains stable under conservative implementation assumptions. As detailed in Supplementary Table S2, the digital screening retains its cost-effectiveness even in the conservative real-world scenario, incorporating substantial digital attrition(45), and reduced clinical effectiveness, yielding an ICER of CHF 4,305 per QALY(53).

#### Policy implications on incentive design

Our analysis identifies digital attrition as the primary source of value leakage for the payer(45). If reimbursement is structured purely as a fee-for-service payment for the digital screening (CHF 40.00), the payer assumes the full financial risk of non-conversion, potentially financing a high volume of digital interactions that fail to translate into clinical value(18).

To solve this misalignment of risks and incentives, this study supports the implementation of a pathway-oriented reimbursement model, potentially piloted under Swiss Federal Health Insurance Act (KVG/LaMal) Article 56 (Experimental Articles) (63). Rather than reimbursing the digital risk biomarker as a standalone service, a pathway-oriented conditional reimbursement contract(64) could combine a reduced upfront payment for the screening event with a performance-contingent component triggered by completed confirmatory blood glucose tests recorded in claims data(1,14).

Such a reimbursement structure would shift the operational risk associated with post-screening digital attrition from the insurer to the digital provider, thereby aligning incentives to facilitate confirmed clinical follow-up rather than maximizing user acquisition alone(15). By linking remuneration to downstream diagnostic completion, developers would be incentivized to invest in implementation features that support real-world care pathways (e.g., appointment facilitation, support medical practice assistants to handle the confirmatory testing or intervention adherence). However, these contract arrangements were not explicitly modeled in the economic analysis and should therefore be interpreted as policy-relevant implications derived from the results, rather than as evaluated outcomes(23).

### 4.2 Strengths and limitations

A primary strength of this analysis is the application of an early HTA framework(15,16) that defines the economic viability frontier during the pre-market phase to guide payers, developers and investors with precise performance targeting (sensitivity, specificity, screening cost, etc.)(17). Robustness was ensured via the CHEERS 2022 standards(23) and a probabilistic Markov complaint model that addresses the efficacy-effectiveness gap. By incorporating a conservative digital attrition parameter, the model accounts for the behavioral friction between digital risk detection and clinical confirmatory action, preventing the overestimation of diagnostic yield common in the digital health literature(18).

Several limitations warrant consideration. First, the parametric uncertainty regarding diagnostic accuracy remains substantial. The base-case sensitivity and specificity (70%) are derived from emerging validation studies rather than large-scale prospective trials. Second, the analysis adopts a statutory health insurer perspective, excluding indirect societal costs such as lost productivity and informal care. Given that T2D is a leading cause of workforce exit, payer perspective likely underestimates the total societal return on investment. Third, while the model parameters were calibrated against Swiss epidemiological data, the core transition logic extrapolates short-term trial data from other regions (DPP/ADDITION-Europe) to a lifetime horizon, a common limitation in chronic disease modeling. We addressed this by applying conservative discounting to the preventive intervention effect to avoid overstating the long-term health and economic benefits.

Future research might include societal benefit, and Swiss health utility weights to optimize the model parameters. Refining the model by separating clinical trajectories, such as cardiovascular and microvascular outcomes, could better capture early detection benefits. Moreover, future economic assessments need to consider indirect peripheral costs, including the psychological burden of false positives and the time patients spend using digital screening tools. It’s also important to recognize that ultimately false positives can diminish trust in digital health technologies and lead to diagnostic anxiety.

## 5. Conclusion

Under conservative assumptions regarding diagnostic performance and follow-up adherence, digital screening for T2D is likely to be a cost-effective use of healthcare resources in Switzerland. The digital screening consistently falls within commonly used cost-effectiveness benchmarks(65) and maintains a high probability of cost-effectiveness across extensive sensitivity analyses, indicating robustness to key sources of parameter uncertainty. Importantly, the results suggest that economic value in a triage setting is driven less by near-perfect diagnostic accuracy than by keeping screening costs low relative to the downstream costs of undetected disease. To translate modeled efficiency into realized payer value, reimbursement approaches that link digital screening to confirmed clinical diagnosis and care pathways are proposed.

## Data Availability

All data generated or analyzed during this study are included in this published article. Additional source codes and details on economic modeling are available at 10.17605/OSF.IO/5G2YR.

https://osf.io/5g2yr/

## Abbreviations

AI: Artificial Intelligence
BIA: Budget Impact Analysis
BMI: Body Mass Index
CDC: Centers for Disease Control and Prevention
CEA: Cost-Effectiveness Analysis
CEAC: Cost-Effectiveness Acceptability Curve
CHEERS: Consolidated Health Economic Evaluation Reporting Standards
CHF: Confoederatio Helvetica Franc (Swiss Franc)
CrI: Credible Interval
DPP: Diabetes Prevention Program
EVPI: Expected Value of Perfect Information
FSO: Federal Statistical Office (Switzerland)
GP: General Practitioner
HTA: Health Technology Assessment
ICER: Incremental Cost-Effectiveness Ratio
KVG: Krankenversicherungsgesetz (Swiss Federal Health Insurance Act)
LaMal: Loi fédérale sur l’assurance-maladie (French abbreviation for KVG)
PSA: Probabilistic Sensitivity Analysis
QALY: Quality-Adjusted Life Year
Se: Sensitivity
Sp: Specificity
T2D: Type 2 Diabetes
TARDOC: Tarifsystème Ambulant (New Swiss Outpatient Tariff Structure)
TARMED: Tarif Médical (Past Swiss Outpatient Tariff Structure)
WTP: Willingness-To-Pay

## Transparency

### Declaration of Financial/Other Relationships

W.M., V.B., M.F., Q.J., E.F., T.K., and M.J. are affiliated with the Centre for Digital Health Interventions (CDHI), a joint initiative of the Institute for Implementation Science in Health Care, University of Zürich; the Department of Management, Technology, and Economics at the Swiss Federal Institute of Technology in Zürich; and the Institute of Technology Management and School of Medicine at the University of St Gallen. CDHI is funded in part by CSS, a Swiss health insurer, MavieNext, an Austrian healthcare provider, and the Swiss investor MTIP. E.F. and T.K. were co-founders of Pathmate Technologies, a university spin-off company that creates and delivers digital clinical pathways. However, T.K. has no shares since 2023 and had never a formal role with Pathmate Technologies. Moreover, neither Pathmate Technologies, MavieNext, nor MTIP was involved in this research. B.W., S.B., and M.B. declare no competing interests.

### Author Contributions

W.M. conceived the study, designed the financial model, performed the analysis, and drafted the original manuscript. B.W., S.B., and M.B. reviewed the clinical validity of model parameters and assumptions. V.B., M.F., and Q.J. contributed to data interpretation and critically revised the manuscript for intellectual content. E.F., T.K., and M.J. supervised the study, provided administrative support, and revised the final manuscript. All authors read and approved the final manuscript for publication.

## Acknowledgements

The authors thank Dr. Irene Salvi (School of Medicine, University of St. Gallen) and colleagues at the Centre for Digital Health Interventions for their feedback on earlier drafts of this work. The authors acknowledge the use of large language models (Grammarly, ChatGPT, Gemini) for language refinement and editing of the manuscript text; all content was reviewed and verified by the authors for accuracy and integrity.

## Availability of data and materials

All data generated or analyzed during this study are included in this published article. Additional source codes and details on economic modeling are available at <u>10.17605/OSF.IO/5G2YR</u>.

## Ethics approval and consent to participate

This study was granted formal approval and a letter of exemption by the ethics committee of the University of St.Gallen, in accordance with institutional guidelines, on 3 February 2026.

