## Supplement material for "Early health technology assessment of digital diabetes screening in Switzerland: cost-effectiveness and budget impact analyses"

**Supplementary Material**

***S1. Markov Trace***

To demonstrate internal validity, Table S1 presents the hypothetical cohort's trajectory through the health states over the 40-year horizon. The results confirm logical disease progression, with the "Healthy" proportion declining over time as individuals transition to "Prediabetes," "Diabetes," or "Death," consistent with natural history assumptions. The values below represent the percentage of each state's surviving cohort at the end of the simulation cycle.

Table S1. Markov chain cohort trace (intervention arm)

| Cycle (year) | Healthy (no diabetes) | Prediabetes | Undiagnosed T2D | Diagnosed T2D | Complication | Death (cumulative) |
|---|---|---|---|---|---|---|
| Year 0 | 60.80% | 27.20% | 3.30% | 8.70% | 0.00% | 0.00% |
| Year 5 | 52.70% | 27.00% | 3.20% | 12.00% | 3.10% | 1.90% |
| Year 10 | 45.80% | 25.80% | 4.00% | 13.10% | 4.70% | 6.50% |
| Year 20 | 34.50% | 21.70% | 4.70% | 12.90% | 6.20% | 20.00% |
| Year 40 | 16.50% | 11.30% | 2.90% | 7.10% | 4.70% | 57.40% |

### *S2. Extended Scenario and Subgroup Analyses*

To assess the robustness of the findings across heterogeneous populations and implementation conditions, stratified analyses were conducted. This analysis explores the economic impact of deviations from ideal trial conditions, specifically modeling the degradation of intervention effectiveness and screening uptake due to real-world systemic friction.

Table S2. Implementation feasibility scenario analysis

| Scenario | Operational description | Parameter calibration | ICER | Δ vs Base | Economic interpretation |
|---|---|---|---|---|---|
| Base case | Idealized trial conditions | Uptake: 30% (1.0×) Effectiveness: 100% $p_{prog}$: 0.0300 | 2,912 | - | Highly Cost-Effective (Dominates standard of care if WTP > CHF 5k) |
| Realistic ramp-up | Year 1 implementation friction; moderate attrition | Uptake: 22.5% (0.75×) Effectiveness: 85% $p_{prog}$: 0.0345 | 4,747 | +63.0% | Robustly Cost-Effective (Remains well below CHF 50k threshold) |
| Pessimistic | High systemic barriers; low uptake; poor adherence | Uptake: 12.0% (0.40×) Effectiveness: 65% $p_{prog}$: 0.0405 | 4,305 | +47.8% | Robustly Cost-Effective (Efficiency driven by low screening cost) |
| Optimistic | Optimized care pathway; enhanced engagement | Uptake: 28.5% (0.95×) Effectiveness: 105% $p_{prog}$: 0.0285 | 2,721 | –6.50% | Efficiency Gain (Marginal value of optimization is low) |

Note: Δ denotes the percentage change in the incremental cost-effectiveness ratio (ICER) relative to the Base Case. Implementation delays (1–2 years) were conceptually applied to the friction scenarios to inform the "Realistic" and "Pessimistic" parameter selection, but are not explicitly modeled as time-lags in the ICER calculation.

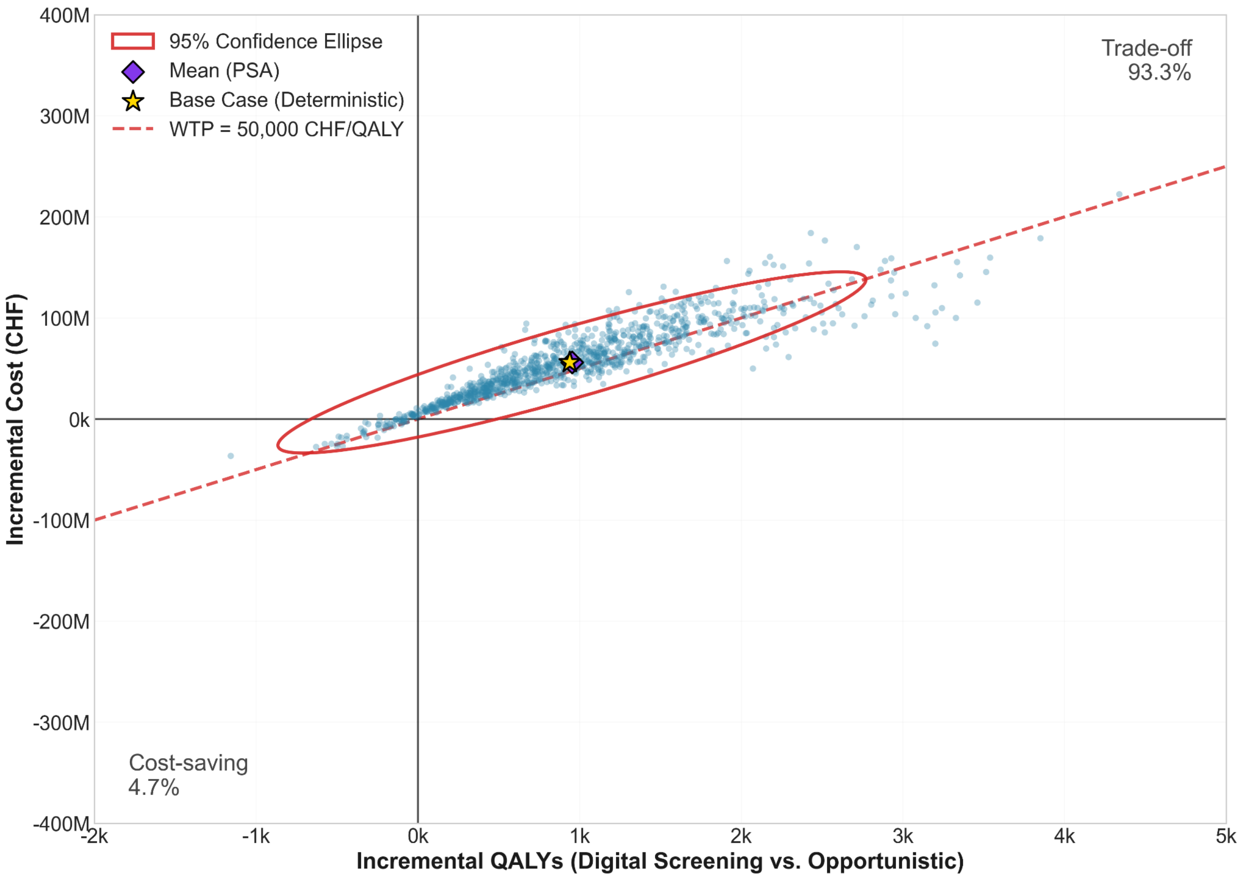


Figure S1. Cost-effectiveness plane averaging ICER of 59,678 CHF/QALY (5-year horizon)

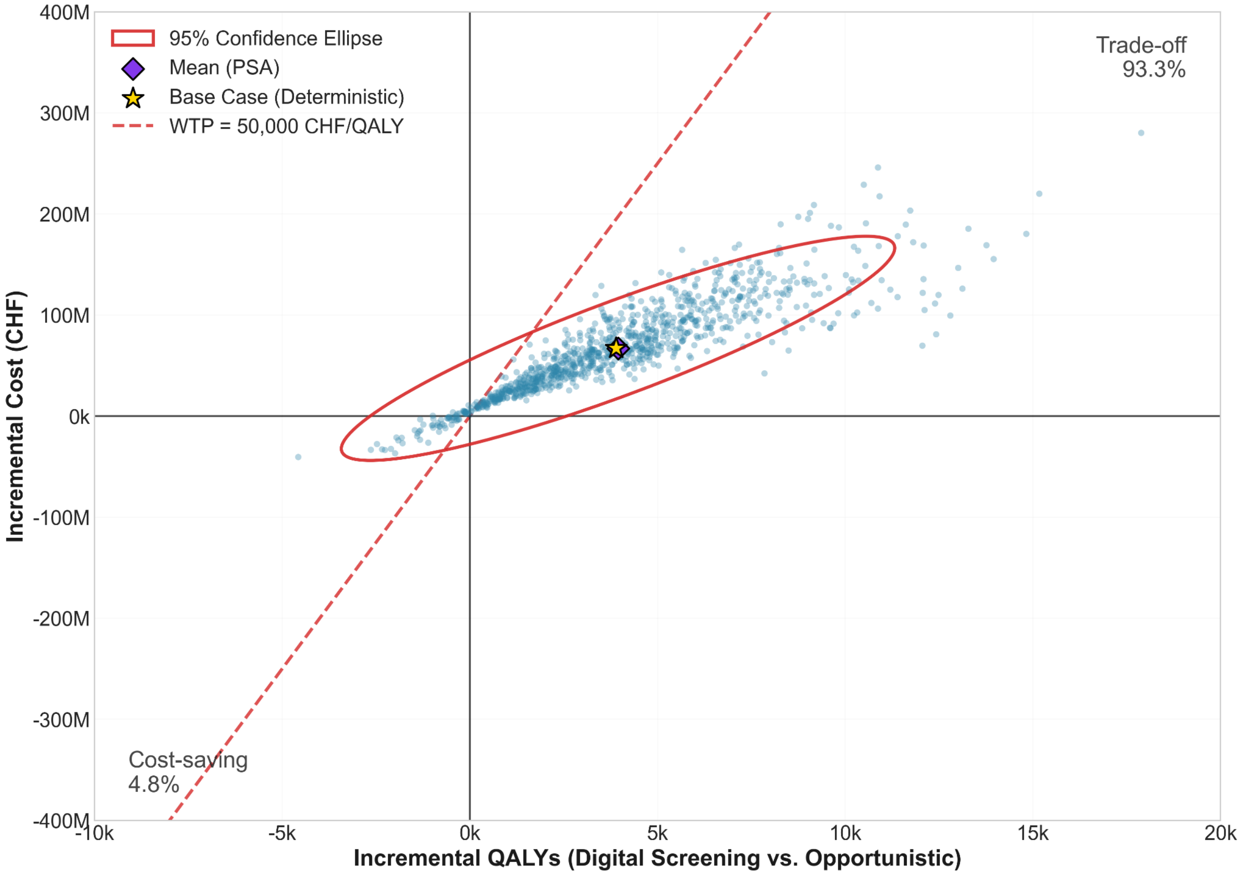


Figure S2. Cost-effectiveness plane averaging ICER of 17,252 CHF/QALY (10-year horizon)

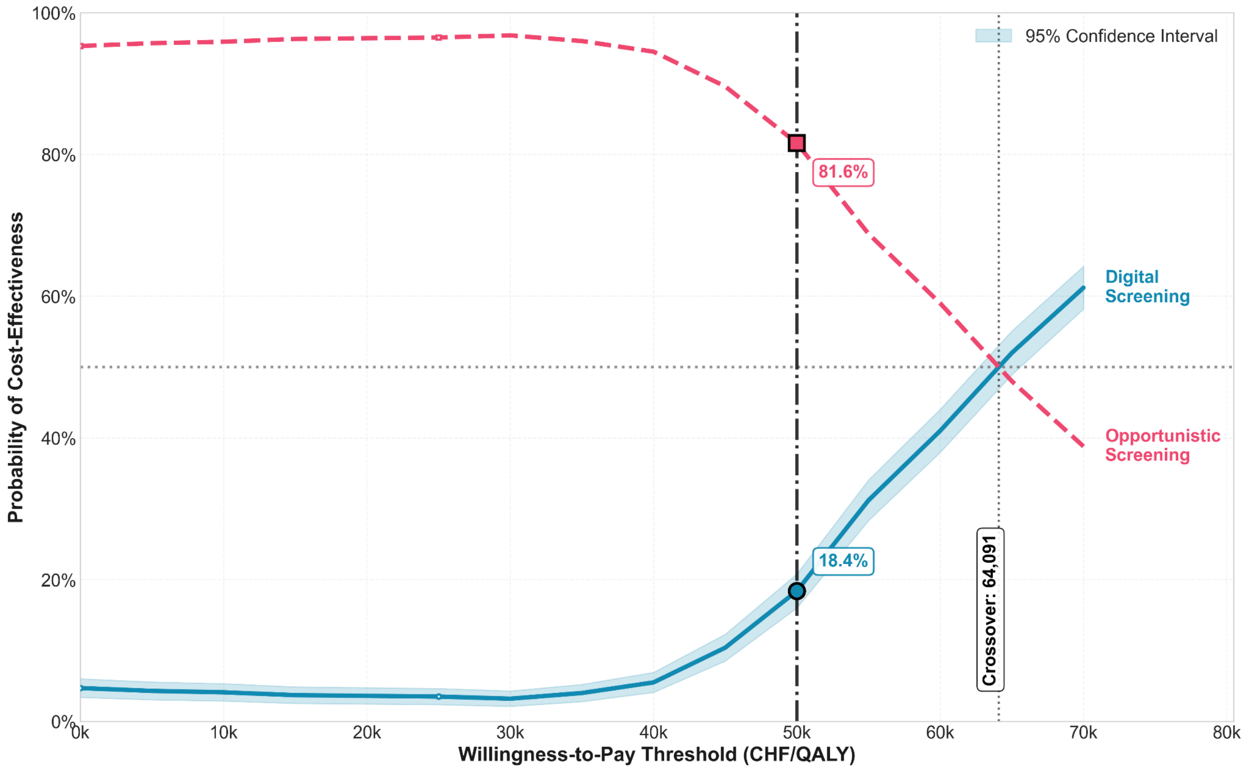


Figure S3. Cost-effectiveness acceptability curve (CEAC) digital vs. opportunistic screening (5-year horizon)

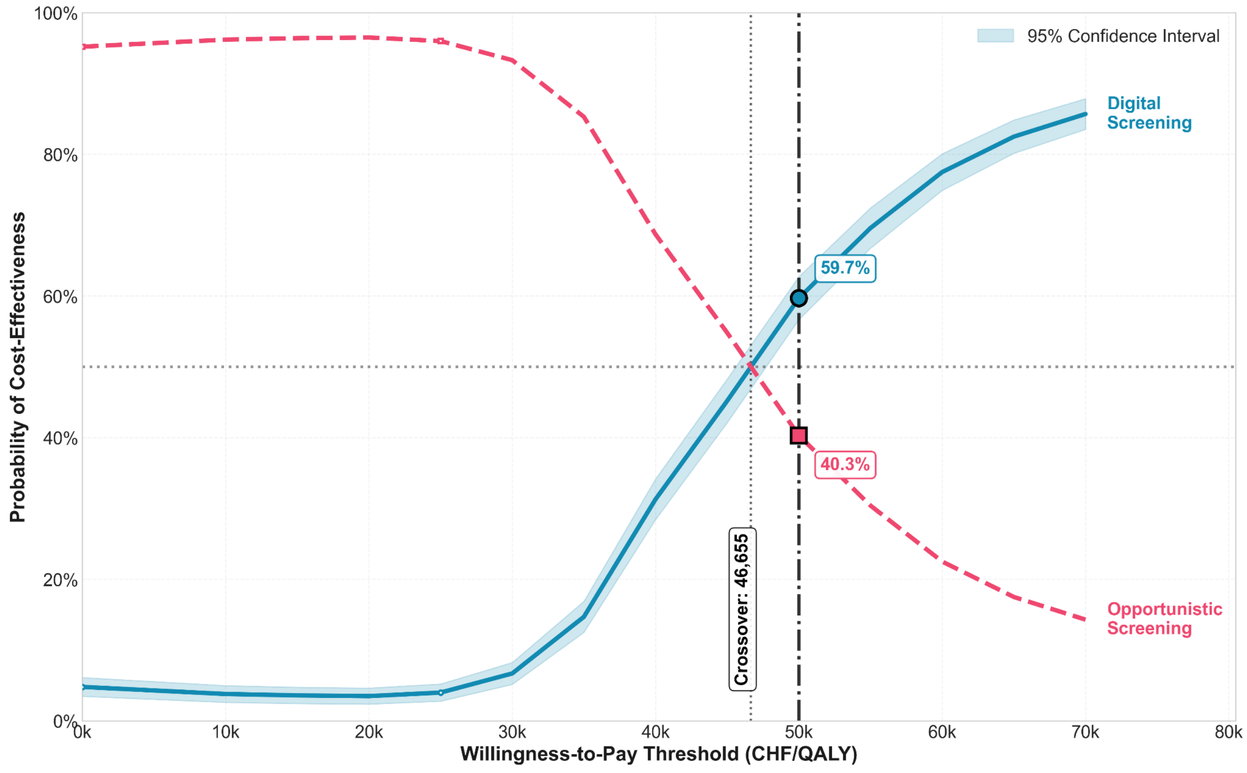


Figure S4. Cost-effectiveness acceptability curve (CEAC) digital vs. opportunistic screening (6-year horizon)

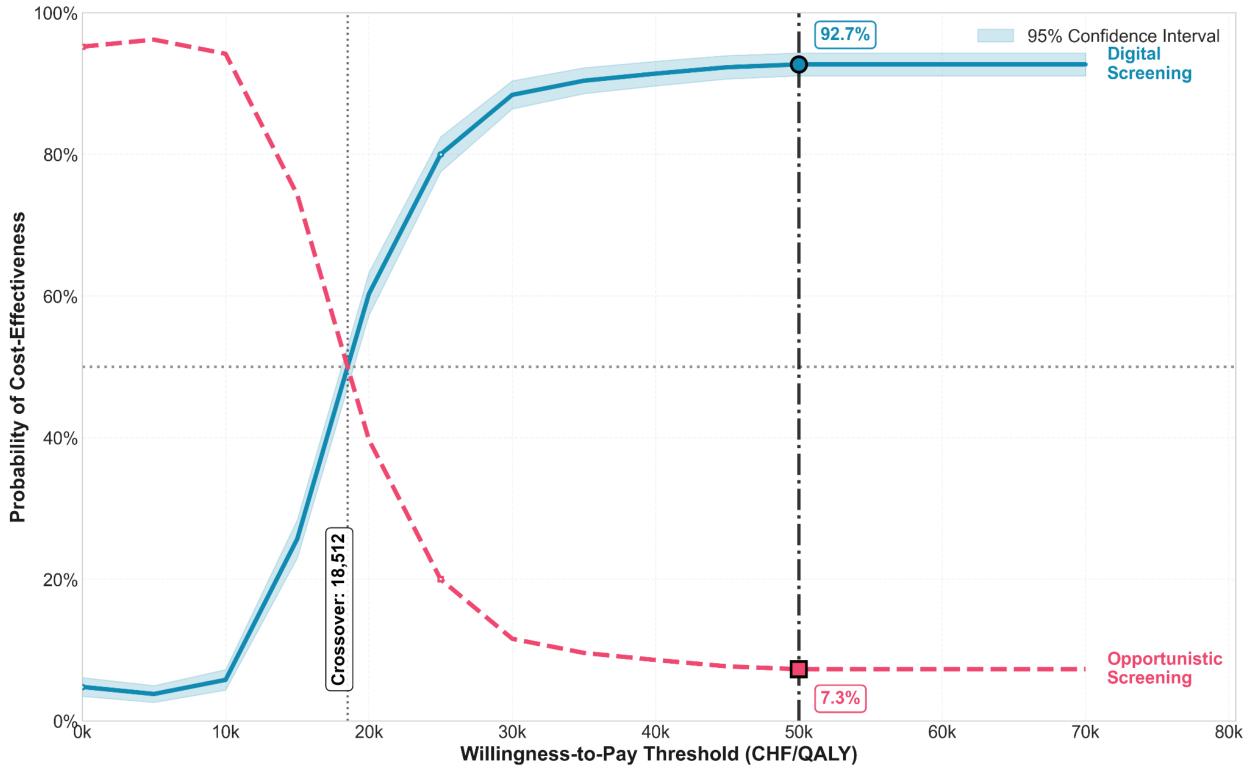


Figure S5. Cost-effectiveness acceptability curve (CEAC) digital vs. opportunistic screening (10-year horizon)

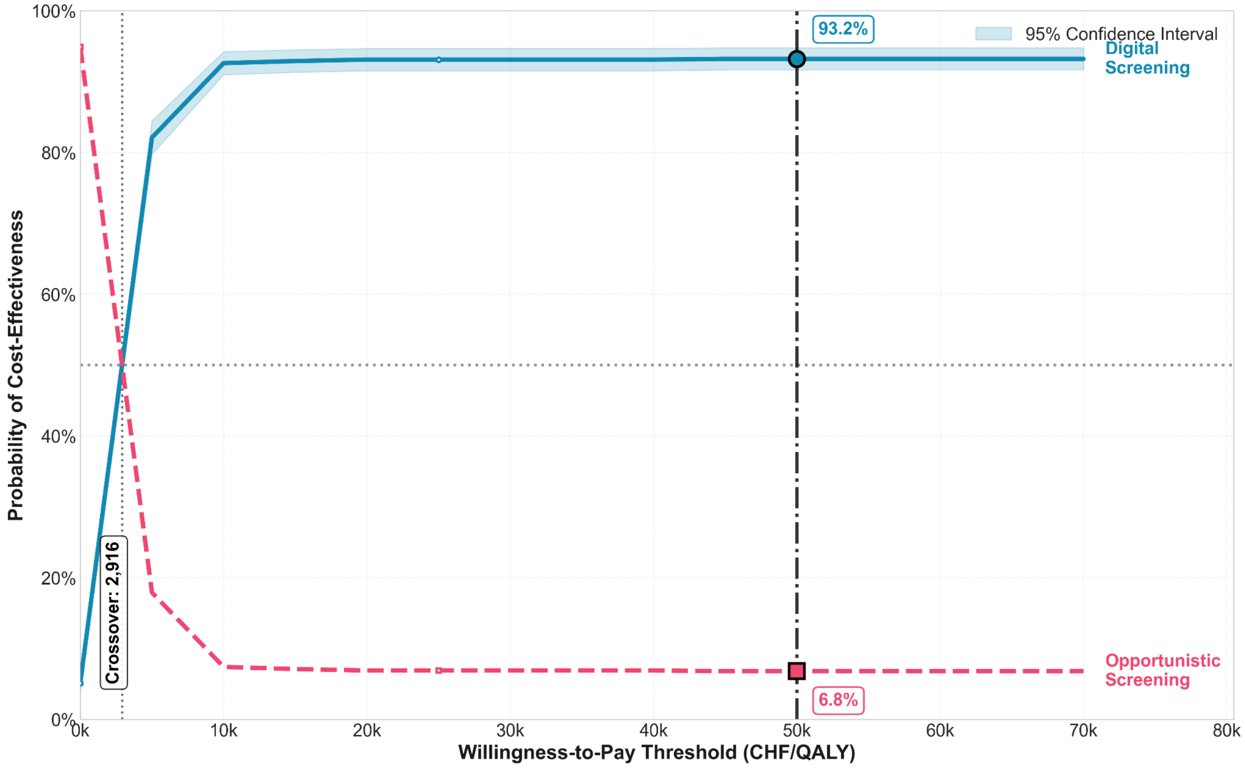


Figure S6. Cost-effectiveness acceptability curve (CEAC) (40-year horizon)

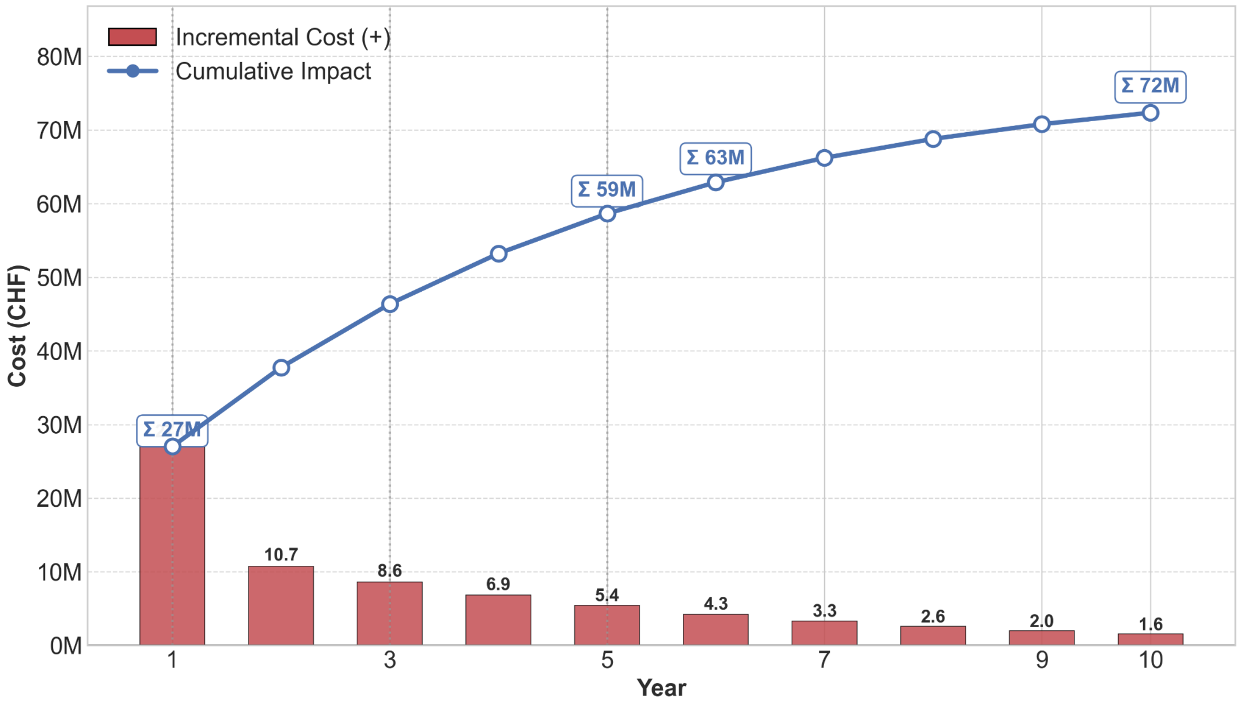


Figure S7. Budget impact by cost decomposition analysis (10-year horizon)

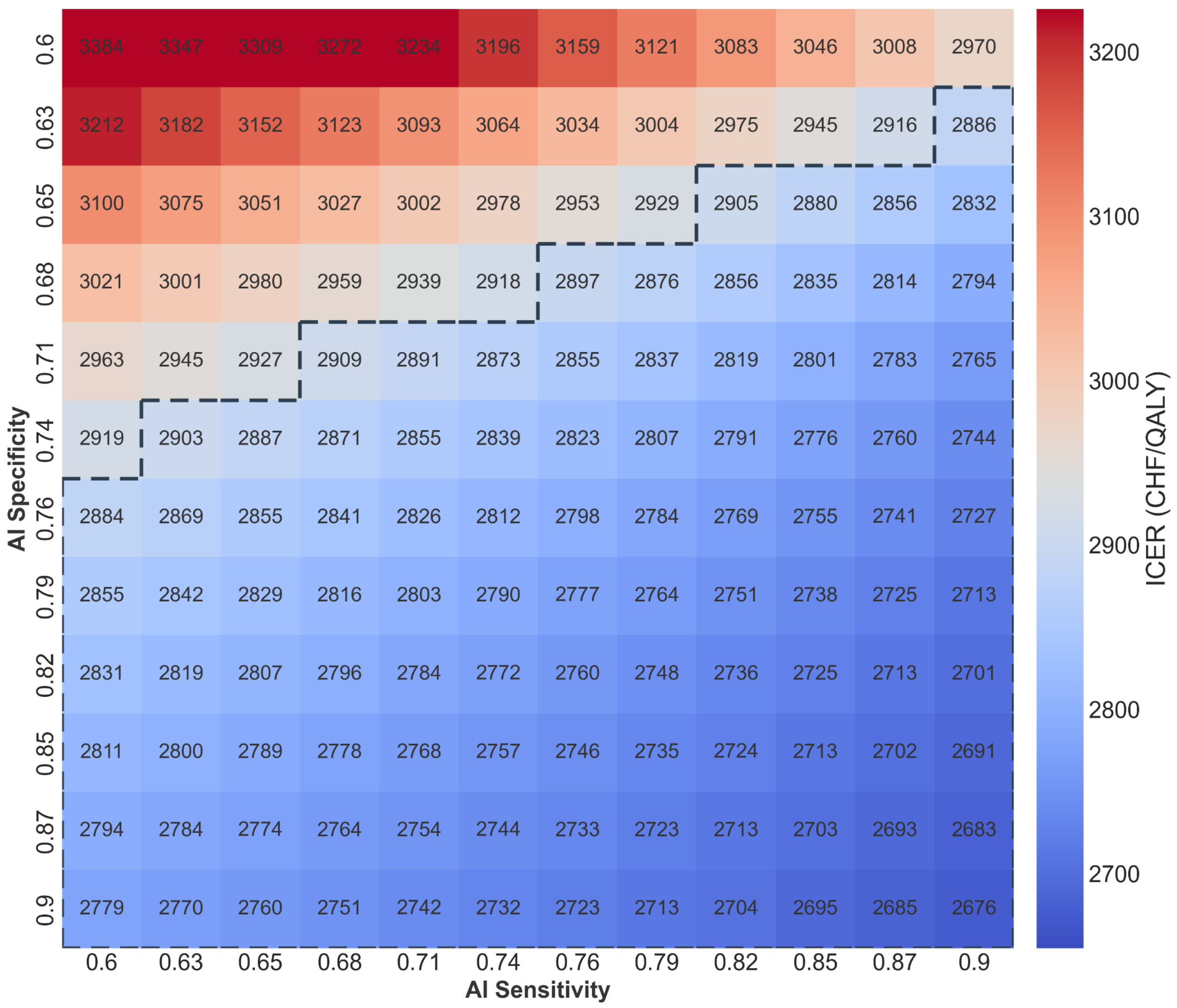


Figure S8. Threshold heatmap of ICER by sensitivity and specificity

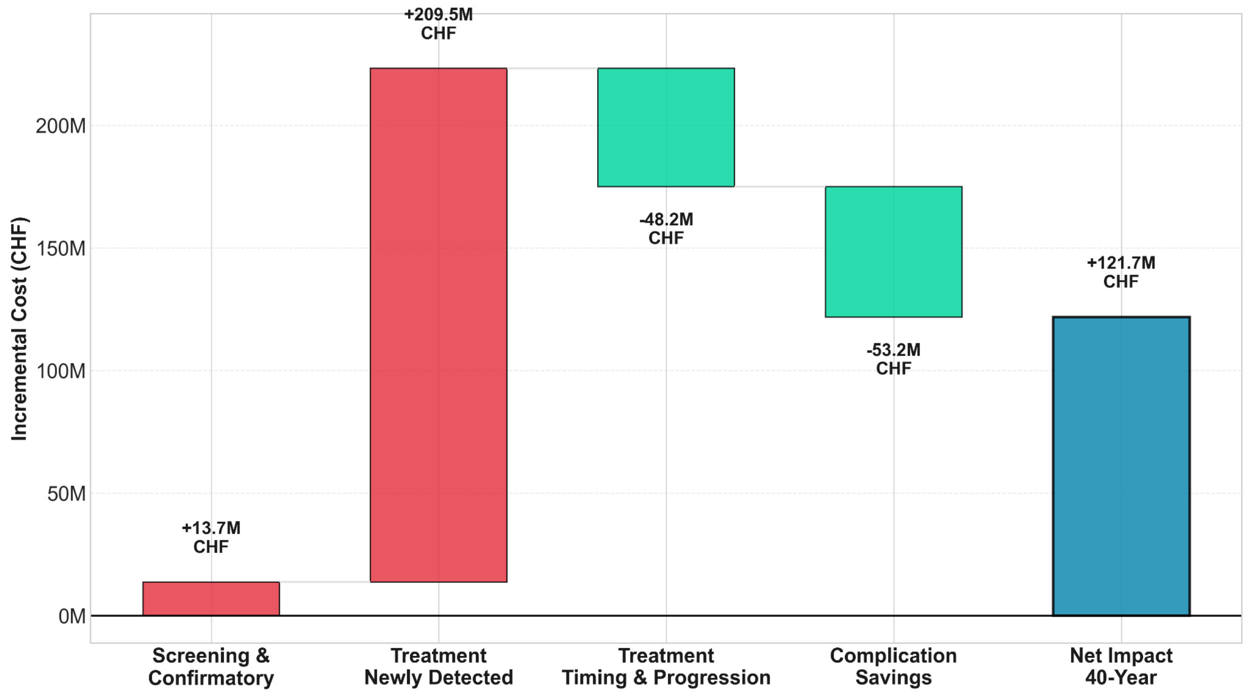


Figure S9. Budget impact by cost decomposition analysis (40-year horizon)
